# Functional gradient perturbation in Wilson disease correlates with structural lesions and transcriptomic specializations

**DOI:** 10.1101/2023.06.19.23291569

**Authors:** Sheng Hu, Chuanfu Li, Yanming Wang, Taohua Wei, Xiaoxiao Wang, Ting Dong, Yulong Yang, Yufeng Ding, Bensheng Qiu, Wenming Yang

## Abstract

Functional dysregulations in multiple regions are caused by excessive copper deposition in the brain for Wilson disease (WD). While the biological mechanism of these dysregulations was thought to be the absent or reduced expression of the ATP7B protein in the liver, mechanisms for such gene impacting brain function remain unexplored. Here, we used a large cohort of resting-state fMRI data (105 WD patients and 93 healthy controls) to derive the functional connectome gradient, and its WD-related alterations were further evaluated. Then, we used Neurosynth, clinical data, and whole-brain gene expression to examine the meta-analytic cognitive function, clinical phenotypes, and transcriptional specializations related to WD gradient alterations. In parallel, spatial correlation between gradient and gray matter volume was accessed for both WD patients and healthy controls. Compared to controls, WD patients exhibited principal gradient alterations in both global and system levels and regional alterations mainly distributed in the sensorimotor, visual, ventral attention, subcortical, and default mode networks. Meta-analytic terms and clinical characters showed the correlations of these gradient alterations in motor-related processing, higher-order cognition, neurological symptom, and age. Results of spatial correlation revealed structure-function decoupling in multiple networks, especially in subcortical and visual networks. Within the cortex, the gradient alterations derived transcriptional specializations of WD that mainly display properties indicative of ion homeostasis, neural development, and motor controls. Within the subcortical regions, we for the first time characterized the role of the ATP7B gene impacting subcortical function. Transcriptional specializations of WD within both cortex and subcortical regions were also associated with neurological and psychiatric disorders, explaining the mechanism underlying complex clinical symptoms from the biological level for WD. In addition, we further illustrated that structural lesion and gradient perturbation shared similar transcriptional specializations in both cortex and subcortical regions for WD. These findings bridged functional gradient perturbation to structural lesions and transcriptional profiles in WD, possibly promoting our understanding of the neurobiological underpinnings underlying the emergence of complex neurological and psychiatric phenotypes.

## INTRODUCTION

Wilson disease (WD) is an autosomal recessive disorder of copper metabolism caused by the lack of biliary copper excretion^1^. The mutation of the ATP7B gene which is most highly expressed in the liver results in the absent and reduced expression of the ATP7B protein, identifying as causative of copper dysmetabolism^2^. Excessive copper accumulation in the liver and brain ultimately leads to neurological and psychiatric phenotypes in WD patients^3^. Animal model of WD, however, has determined that the ATP7B gene is also expressed in neuronal cells of the hippocampus, olfactory bulbs, cerebellum, brainstem, and cerebral cortex. The concurrent high levels of copper in these regions further implicated that dysfunctional ATP7B protein is related to local copper accumulation and the cerebral manifestations of WD^4^.

The toxic effect of copper results in damaged astrocytes, neurons, and oligodendrocytes, which gradually build up structural lesions, especially in the subcortical regions, including the basal ganglia, thalamus, and brainstem^5,6^. Of note, functional MRI studies revealing functional abnormities across the entire brain suggested that a structure-function decoupling may be the leading cause for these abnormities in WD^7,8^. Further, the defective cortical function may be due to dysfunction of cortio-subcortical pathways, leading to neurological and psychiatric symptoms in WD^9,10^. However, the neurobiological mechanisms underlying the brain dysfunction remain to be elucidated. Given that gene has an essential influence on brain function^11^, brain dysfunction of WD may also be conceivable to shape by the specific mutated gene, such as ATP7B. Moreover, in consideration of the variability of gene expression profile across the entire brain, such as the cortex and subcortical regions^12^, their relation to the WD topology of dysfunction may provide insights into the neurobiological pathways contributing to the neurological pathogenesis of WD.

Brain hierarchical theories uncovered that functional connectome patterns are governed by the graded macroscale axe that spans the primary sensorimotor areas to transmodal regions of the default mode network (DMN)^13^. Perturbation in these patterns shapes the clinical phenotypes, such as perception, cognition, and neurological disorder, in multiple neurodegenerative diseases^14,15^. Furthermore, the macroscale axe of the cortex also is intrinsic in gene expression and microstructural topography, indicating that the functional basis strongly depends on the intact gene expression and microstructural profiles^12^. Although many studies revealed alterations in functional connectivity involving basal ganglia, sensorimotor, and DMN regions in patients with WD^7,8,16^, no studies reported whether and how these alterations are affected by structural and transcriptomic vulnerability. Therefore, if WD patients exhibit perturbation in the functional hierarchy, these abnormalities might be associated with gene expression and structural topography. The illustration of such alterations would provide complementary insights into the structural and molecular genetic underpinnings of dysfunctional hierarchy in WD.

Functional connectome patterns are facilitated by decomposing the functional connectivity data into different gradients to capture the topography of the connectome^17^. Such a principal functional gradient is presented in the macroscale organization of functional brain networks, reflecting the hierarchical macroscale axe^13^. This study aimed to investigate the structural and genetic underpinnings of functional hierarchical perturbation in WD. For this purpose, we mapped the functional gradient using functional MRI and determined the perturbation in the hierarchical macroscale axe for WD. Then, the gray volume was mapped based on 3D high-resolution MRI to apply to a function-structure coupling analysis, revealing whether function-structure is decoupled in WD. Meanwhile, we calculated the spatial correlation of connectome gradient perturbation with whole-brain gene expression data from the Allen Human Brain Atlas^18^ (AHBA) and further examined whether structural lesions and gradient perturbation shared similar transcriptional specializations. Gene enrichment analysis was finally used to probe transcriptomic associations for known pathogenic WD variants, as well as other neurological and psychiatric disorders.

## MATERIALS AND METHODS

### Participants

We studied a larger cohort of 105 WD patients (65 males, mean ± SD age = 27.29 ± 4.02 years) and 93 healthy controls (60 males, mean ± SD age = 24.48 ± 2.26 years) collected from the First Affiliated Hospital of Anhui University of Chinese Medicine. All patients met the diagnostic criteria, with a ceruloplasmin concentration < 0.1 g/L, a 24-h urinary copper excretion >100 μg, and the presence of a K-F ring on slit-lamp examination. Unified Wilson’s Disease Rating Scale neurological examination (UWDRS-N) subscores were recorded as a measure of neurological disorder severity^19^. Exclusion criterium for WD patients was diagnosed with other diseases. Normal individuals with a history of mental health problems or other serious diseases were excluded from this study. Ethics committee of the First Affiliated Hospital of Anhui University of Chinese Medicine gave ethical approval for this work, and written informed consent was obtained from all participants. Demographic and clinical information are described in **Table 1**.

**Table 1.** Demographic and clinical characteristics of the WD and HCs

|  | WD<br>n = 105 | HCS<br>n = 93 | P-value |
| --- | --- | --- | --- |
| Age mean (SD), years | 27.23 ± 8.30 | 24.48 ± 2.26 | 0.0023 |
| Males, n (%) | 59 (60%) | 60 (64%) | - |
| Education mean (SD), years | 11.97 ± 3.39 | 15.42 ± 4.06 | < 0.001 |
| Disease duration mean (SD), year | 9.13 ± 6.67 | - | - |
| UWDRS-N score mean (SD) | 10.06 ± 12.64 | - | - |
| Ceruloplasmin mean (SD), g/L | 0.041 ± 0.04 | - | - |
| 24h urinary copper excretion mean (SD), ug | 850.85 ± 588.56 | - | - |

### Data acquisition and preprocessing

Data were obtained at the First Affiliated Hospital of AUCM using a GE MR750 scanner. Resting-state functional MRI (R-fMRI) was acquired using a gradient-echo single-shot echo-planar imaging sequence, with the following parameter settings: repetition time (TR), 2000 ms; echo time, 30 ms; flip angle, 90°; matrix size, 64 × 64; field of view, 220 mm × 220 mm; resolution of the axial slice, 3.4375×3.4375 mm^2^; slice thickness, 3 mm. A total of 185 volumes were acquired from each individual with 36 slices per volume. T1-weighted images were collected using a T1-3D BRAVO sequence: TR, 8.16 ms; echo time, 3.18 ms; flip angle, 12°; matrix size, 256 × 256; field of view, 256 mm × 256 mm; voxel size, 1 mm^3^; slice thickness, 1 mm. R-fMRI data were preprocessed using a standard pipeline, including slice timing, head motion correction, normalization, and bandpass, described in **Supplementary Materials**.

### Gray matter volume estimation

Gray matter (GM) volume measurements were performed using a voxel-based morphometric (VBM) approach^20^ that is embedded in the FSL-VBM toolbox. Briefly, the gray matter was first segmented from the T1-weighted images, followed by normalized to MNI152 standard space. The normalized images were further applied to create a study-specific GM template. All native GM images were subsequently registered to the template and modulated for contraction. The modulated GM images were finally smoothed using an isotropic Gaussian kernel with a sigma of 3 mm. GM volume differences between WD and HCs were evaluated using permutation-based non-parametric testing with random permutations. The statistical significance was set as P < 0.001, using a threshold-free cluster enhancement (TFCE) approach with family wise-error (FWE) correction for multiple comparisons^21^.

### Preprocessing gene expression data

The microarray gene expression data from six donors (mean age: 42.5 years, 5 males and 1 female) were downloaded from the Allen Human Brain Atlas (AHBA, http://human.brain-map.org/)^22^. The six post-mortem brains were segmented into 3702 spatially distinct tissue samples, with each tissue sample normalized to Montreal Neurological Institute (MNI) coordinate space, and indexed with expression levels for more than 20,000 genes from at least two probes^22^. We respectively conducted gene expression analysis within the cortical and subcortical regions due to the large transcriptional differences between the two main regions^23^. The standard preprocessed pipeline of the gene expression microarray data was performed in the current study, including mapping the tissue samples onto a cortical parcellation with 360 parcels^24^ and a subcortical parcellation with 54 parcels^25^, probe reannotation and selection, and normalization across donors^18^.

### Functional connectome gradient analysis

We constructed individual functional connectomes at the voxel level. To reduce the computational complexity, we resample the preprocessed R-fMRI data to a 4-mm isotropic resolution. For each individual, a functional connectivity (FC) matrix was first constructed by performing the Pearson correlation analysis between each pair of gray matter nodes (20208 voxels). Then, the FC matrix was further applied to the diffusion map embedding method to calculate the connectome gradient^17,26^. Specifically, the FC matrix was thresholded to retain the top 10% connections of each node, and the cosine similarity between each pair of nodes was computed. Further, the similarity matrix was scaled into a normalized angle matrix to avoid the negative values^27,28^. The diffusion map embedding approach was finally applied to identify gradient components that explain most functional connectome variances. Following the previous recommendation, we set the manifold learning parameter α = 0.5 in the diffusion process^29^. Given that the principal gradient to an extreme represents the macroscale axe, we primarily focused on perturbation in the principal gradients of WD^13^. The case-control differences between WD and HCs in the connectome gradient were evaluated by a two-sample t-test with age, sex, and education as covariates. The false discovery rate (FDR) was used to achieve multiple comparison corrections at the voxel level. The statistically significant threshold was set to *q* < 0.05. In addition, the whole-brain voxels were assigned to eight systems according to a cortical atlas with 7 parcels and a Harvard-Oxford probabilistic subcortical atlas. The system-based differences in connectome gradient between WD and HC were further examined using a paired two-sample t-test. The findings were further corrected by multiple comparisons using FDR at 0.05.

### Association analysis between meta-analytic cognitive terms and gradients changes in WD

The Neuosynth (https://neurosynth.org/)^30^ was used to evaluate the relationship between the meta-analytic cognitive terms and functional gradients alternations in WD. The thresholded Z-map derived from the between-group comparisons of the gradient was first divided into WD-positive (WD > HCs) and WD-negative (WD < HCs) maps. The ‘decoder’ function in Neurosynth was used to identify the spatial correlations between each map and the meta-analytic map of each term in the database^31^. Finally, the top 30 cognitive terms were selected.

### Correlation analysis between clinical characters and gradients in WD

We examined the relationships between gradients in perturbed regions and clinical characters, such as UWDRS-N, age, ceruloplasmin concentration, and 24-h urinary copper excretion, using partial correlations with age and sex controlled.

### Association analysis between the GM volume and gradients

To investigate whether structure-function is decoupled in WD, the spatial association between averaged GM volume and averaged functional connectome gradients was estimated using spatial correlation analysis. The spatial correlation analyses were performed based on the whole brain and eight systems. For all spatial correlation analyses, findings were corrected using permutation tests (N = 10,000) at *P* = 0.05. The results were further corrected for multiple comparisons using FDR at 0.05. The spatial correlation was also calculated at the subject level. Then, the differences in spatial correlation between WD and HCs across subjects were accessed using a two-sample t-test, and the results were corrected by FDR at 0.05. Finally, using a non-parametric bootstrapping approach (10,000 times), we performed a meditation analysis in which altered gradient was taken as an independent variable, GM volume in altered gradient regions was taken as the mediator, and UWDRS-N was taken as the dependent variable.

### Association analysis between gene expression and WD’s gradients changes

The spatial correlations between-group differences of principal gradient and gene expression were respectively examined in cortical and subcortical regions. Given the high dimensionality of AHBA data, we used partial least squares (PLS) regression^32^, a multivariate linear model, to reveal a group of weighted genes (or PLS components) that best explained the differences of the gradient. Briefly, we first aligned the gene expression data (10028 genes) and between-group difference Z-map of the principal gradient to a cortical atlas^24^ and subcortical atlas^25^. In PLS analysis, the gene expression data was used to identify the linear combinations of genes that best predicted response variables^33^ (Z-map between-group difference of the principal gradient). The statistical significance of the first two PLS components was tested based on 10,000 permutations of the response variables. A bootstrapping approach was finally used to correct the estimation error of the weight of each gene on each PLS component^33^.

### Enrichment analysis

We first ranked the genes according to their bootstrap weights (absolute value). Then, the top 10 percentile of 10028 genes were applied to a web-based gene set analysis toolkit^34^ (https://webgestalt.org) to uncover biological processes enriched in the list of genes. The enrichment ratio is calculated as the number of PLS-derived genes overlapping with each biological process divided by the number of genes expected to overlap by random permutations. Significant enrichment was determined with Bonferroni FDR-corrected *q* < 0.05.

### Specificity analysis

The specificity analysis was to assess whether known genes (ATP7B) causing WD was enriched in the PLS components. We also performed specificity analysis to uncover the enrichment of other neurological and psychiatric risk genes in the PLS components. The disorder-related risk genes, provided by the AHBA (https://help.brain-map.org/display/humanbrain/Documentation), are listed in **Supplementary Table 1**.

We calculated the enrichment ratio (ER) for each PLS component. The ER is defined as the difference between the mean bootstrap weight of the candidate gene and the mean bootstrap weight of the same number of randomly permuted genes, which was further divided by the standard deviation weight of the permuted genes^35^. Significance was determined by the percentile of the bootstrap weight of the candidate genes relative to the bootstrap weights of randomly selected genes from 10,000 permutations. Positive/negative ER of a given condition indicates that the risk genes are expressed to a higher/lower degree relative to the baseline expression level.

### Association analysis between structural changes and PLS components

To investigate whether structural changes and disturbances in connectome gradient shared a similar transcriptional specializations, we calculated the spatial correlations between structural changes and each PLS component in WD. Structural changes were defined as the absolute value of between-group difference of the GM volume in high (PLS-score > 0.06) or low (PLS-score < −0.06) gene expression regions. The spatial correlations were corrected for spatial autocorrelations by using a permutation test (N = 10,000).

## RESULTS

### Perturbation of primary-to-transmodal gradient in WD

The principal gradient explained 12.2 ± 1.8% of the total variance in the connectome across all individuals (WD: 11.8 ± 1.7%, HC: 12.9 ± 1.8%, **Supplementary Fig. 1**), which was organized along graded macroscale axe from the primary visual/sensorimotor (VIS/SMN) areas to DMN (**Fig. 1a**), being consistent with the previous observation of connectome gradients from primary to the transmodal regions in healthy brain^17^. The spatial patterns of principal gradient maps were highly similar between the WD and HCs (Spearman’s R = 0.9734, *P* < 0.0001, spatial autocorrelation corrected by permutation tests, **Supplementary Fig. 2**). The global histogram confirmed that the extreme values were suppressed in WD relative to HCs, while those in the mid-range increased (**Fig 1b**). For each system, we further computed the gradient score differences between group-averaged maps of WD and HCs by using paired t-test across voxel. The WD had a higher gradient score in VIS, SMN, VAN, FPN, and SUB but lower in LIB and DMN (FDR-corrected *q* < 0.05, **Fig. 1c** and **Supplementary Table 2**). The results of the secondary gradient are provided in **Supplementary Figs. 3** and **Supplementary Tables 4**.

**Fig. 1.**
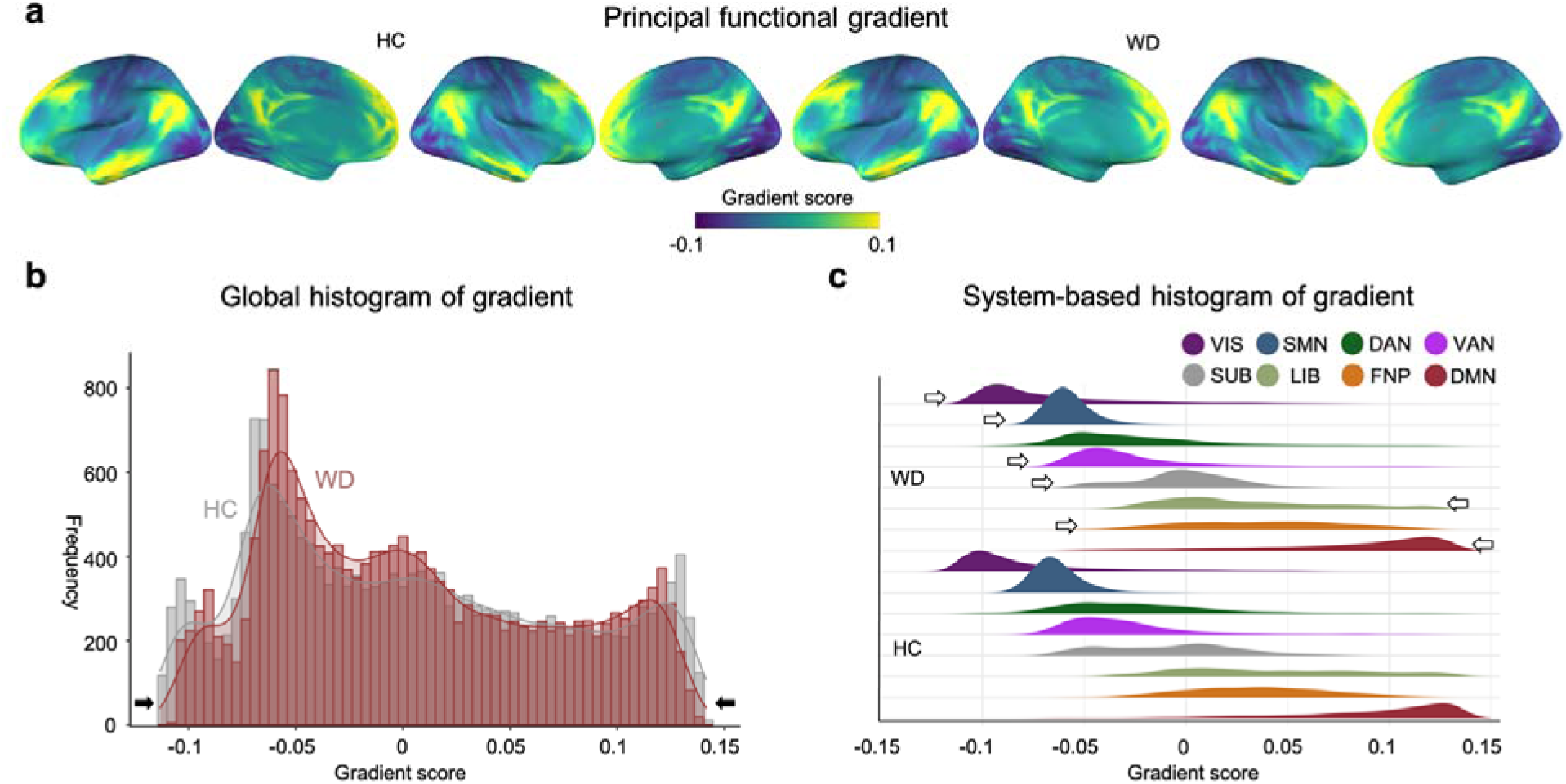
Connectome gradient mapping in WD patients and HCs. **(a)** The principal gradient was organized along a gradual axis from the primary visual/sensorimotor networks to the default mode network. **(b)** Global histogram of the gradient. the extreme gradient values were suppressed in the WD patients relative to those of HCs. **(c)** System-based histogram of the gradient. Arrows indicate the direction of the significant differences between the WD patients and HCs. VIS, visual network; SMN, sensorimotor network; DAN, dorsal attention network; VAN, ventral attention network; SUB, subcortical regions; LIB, limbic network; FPN, frontoparietal network; DMN, default mode network.

### Changes in connectome gradient in WD

The WD showed lower gradient scores mostly in the DMN [medial temporal pole (mTP) and medial prefrontal cortex (mPFC), 70.3%] and LIB (19.4%) but higher scores mainly in the VIS [primary visual cortex (V1), 22.8%], SMN[supplementary motor areas (SMA), 19.9%], VAN (insular, 23.3%) and SUB (striatum, thalamus, 20.6%) than the HCs (absolute *d* = 0.08-0.19, FDR-corrected *q* < 0.05, **Fig. 2a**, **Supplementary Table 3**). For secondary connectome gradients, only one region showed a significantly higher gradient score in WD when compared with HCs (**Supplementary Figs. 4** and **Supplementary Tables 5**.).

**Fig. 2.**
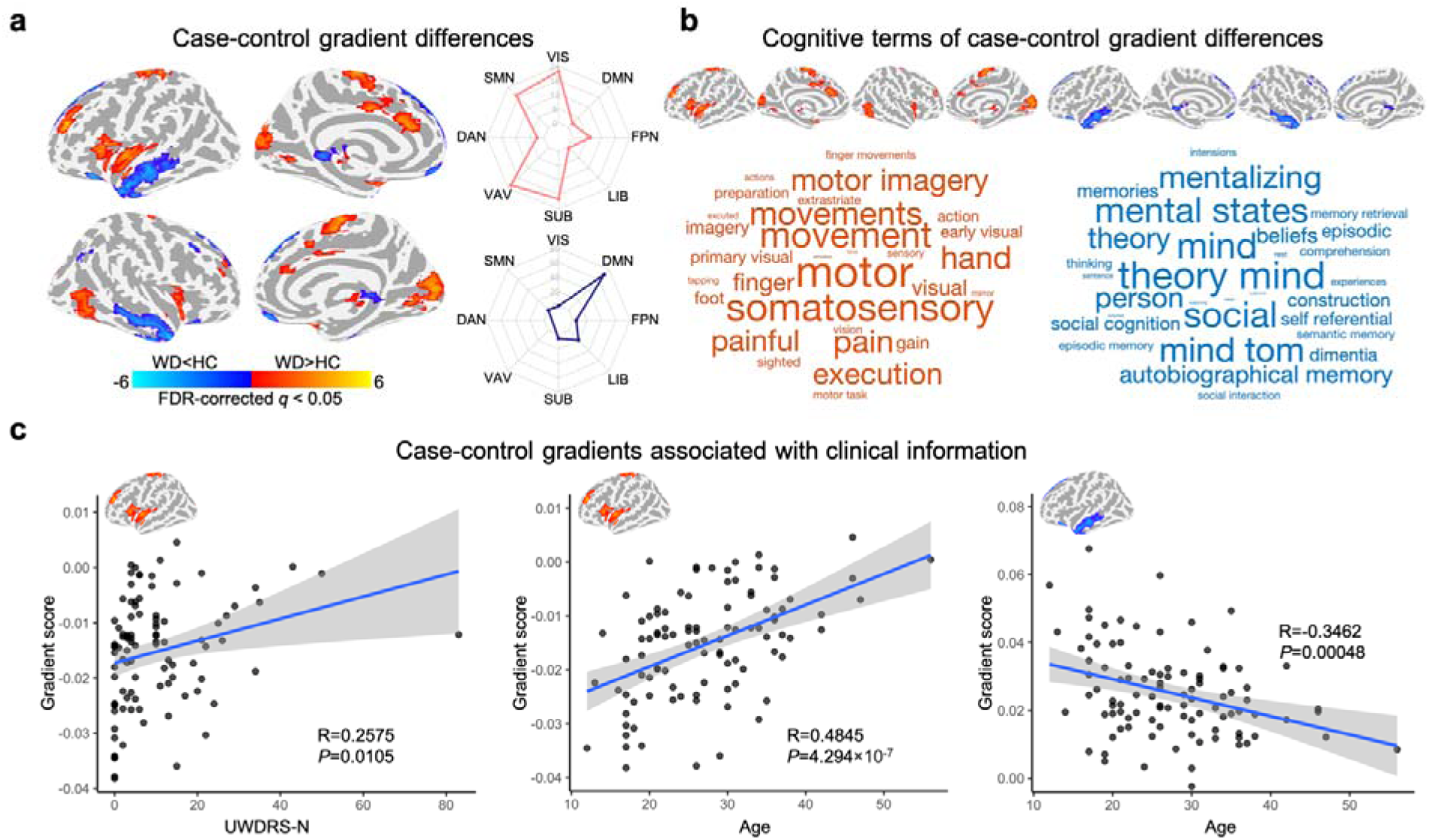
Gradient differences and their associations with cognitive function. **(a)** Voxel-wise statistical comparisons between the WD patients and HCs and distribution of the regional case-control difference in different systems. Higher/lower values in WD are presented as warm/cold colors. The statistical significance level was set as FDR voxel level-corrected *q* < 0.05. **(b)** Word clouds of cognitive functions associated with brain regions that exhibited higher (red) or lower (blue) gradient scores in WD. The font size of a given cognitive term corresponds to the correlation coefficient of the between-group Z-map of the principal gradient with the meta-analytic map of that term generated by Neurosynth. **(c)** Association between regional gradient score and individual UWDRS-N score and age. VIS, visual network; SMN, sensorimotor network; DAN, dorsal attention network; VAN, ventral attention network; SUB, subcortical regions; LIB, limbic network; FPN, frontoparietal network; DMN, default mode network.

### Association between gradient changes and cognitive functions in WD

Higher principal gradient scores in WD were mainly involved in motor-related processes, such as movement, motor imagery, and somatosensory (**Fig2. B and Supplementary Table 6**). Lower principal gradient scores in WD were correlated with higher-order cognitive processes, including mental states, mentalizing, theory mind, and social (**Fig2. B and Supplementary Table 6**).

### Correlations between the principal gradients and clinical characters

The regions with significantly higher gradient scores in WD were positively correlated with UWDRS-N (R = 0.2575, *P* = 0.0105) and age (R = 0.4845, *P* = 4.294 × 10^-7^). The regions with significantly lower gradient scores in WD were negatively correlated with age (R = −0.3462, *P* = 0.00048). No significant correlations were observed between gradient scores with ceruloplasmin concentration, 24-h urinary copper excretion, and disease duration.

### Associations between GM volume and primary-to-transmodal gradient

Evaluating group differences in GM volume between WD and HCs, we found GM volume was significantly reduced in SMN, VIS, DMN, and SUB in WD (**Supplementary Fig. 6 and Supplementary Table 7**). Conversely, significant GM volume was increased in VAN, DAN, LIB, and FPN in WD (**Supplementary Fig. 6 and Supplementary Table 7**).

High spatial correlations between principal gradient and GM volume were observed in VIS, SMN, VAN, SUB, and LIB (**Fig. 3b**) in both WD and HCs, but not in the other systems and the whole brain (**Fig. 3a)**. In the subject level, the spatial correlation was significantly different in the extentive networks, especially in VIS and SUB (**Fig. 3c**), indicating function-structure decoupling in these regions. The spatial correlation between the secondary gradient and GM volume is revealed in **Supplementary Fig 6**.

**Fig. 3.**
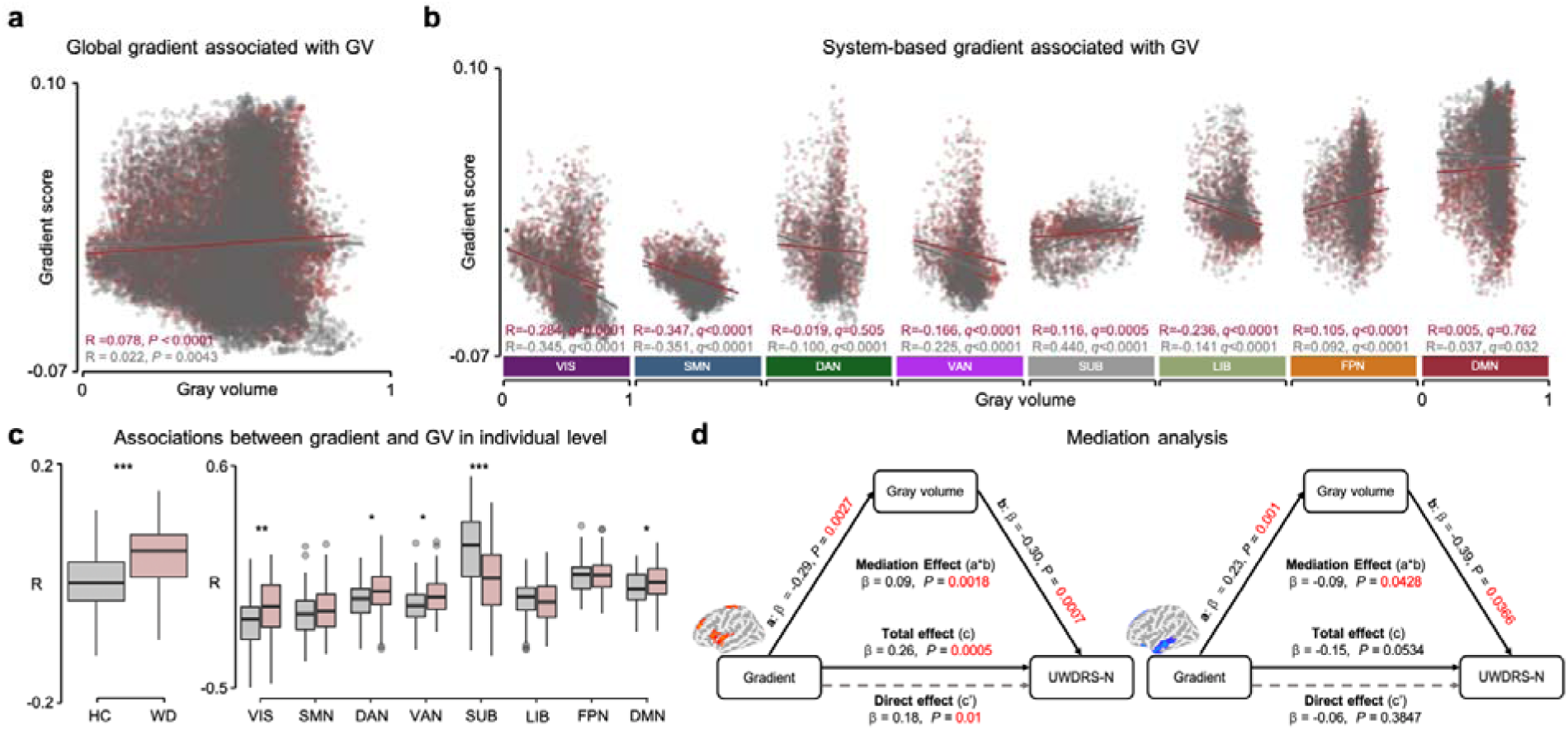
Spatial associations between connectome gradient and GM volume in WD and HC. **(a)** Global association between gradient and GM volume in WD patients and HCs. **(b)** System-based associations between gradient and GM volume in WD patients and HCs. The *P* values were corrected by FDR. **(c)** Global and system-based associations between gradient and GM volume at the individual level and their group differences were measured by a two-sample t-test and corrected by FDR. **(d)** GM volume significantly mediated associations of regional gradients with UWDRS-N score. *, P < 0.05; **, P < 0.01; ***, P < 0.001; GV, GM volume; VIS, visual network; SMN, sensorimotor network; DAN, dorsal attention network; VAN, ventral attention network; SUB, subcortical regions; LIB, limbic network; FPN, frontoparietal network; DMN, default mode network.

We also found that the GM volume in regions with higher (meditation effect: fJ = 0.09, P = 0.0018) and lower (meditation effect: fJ = −0.09, P = 0.0428) gradient scores in WD partially mediated the relationship between gradient and UWDRS-N (**Fig. 3d**).

### Associations between gene expression and WD-related gradient changes

For cortex, two PLS components explained 23% (PLS-1, *P* < 0.001) and 21% (PLS-2, *P* < 0.001) of the covariance between the gradient changes of WD and gene expression (**Supplementary Fig. 7**). PSL-1 represented a gene expression profile with high expression mainly in SMN and VIS but low expression in LIB and DMN (**Fig. 4a**). PLS-2 characterized by high expression in VAN but low expression in FPN (**Fig. 4a**). The gene enrichment analysis shown that PLS-1 involved in regulation at metal ion transport, ion transmembrane transport, and transporter activity (FDR < 0.05, **Fig. 4b**), determining the metal ion homeostasis. By contrast, PLS-2 mainly reflected the muscle system process, circulatory system process, and neuron projection development, as well as regulation at metal ion transport and ion transmembrane transport (FDR < 0.05, **Fig. 4b**). Specificity analysis revealed that PLS-1 and PLS-2 were enriched for the risk genes (**Fig. 4c**), such as Schizophrenia (PLS-1: FDR = 0.0095, ER = −3.05; PLS-2: FDR = 0.0023; ER = −3.37), Depression (PLS-1: FDR = 0.0206, ER = −2.46; PLS-2: FDR = 0.044, ER = 1.90), and Parkinson’s disease (PLS-2: FDR = 0.044, ER = 1.94). The GM volume changes had a significant positive correlation with PLS-1 scores higher than 0.06, indicating that structural impairments and gradient perturbation in cortical regions may be mediated by the similar transcriptional specializations (**Fig. 4d**).

**Fig. 4.**
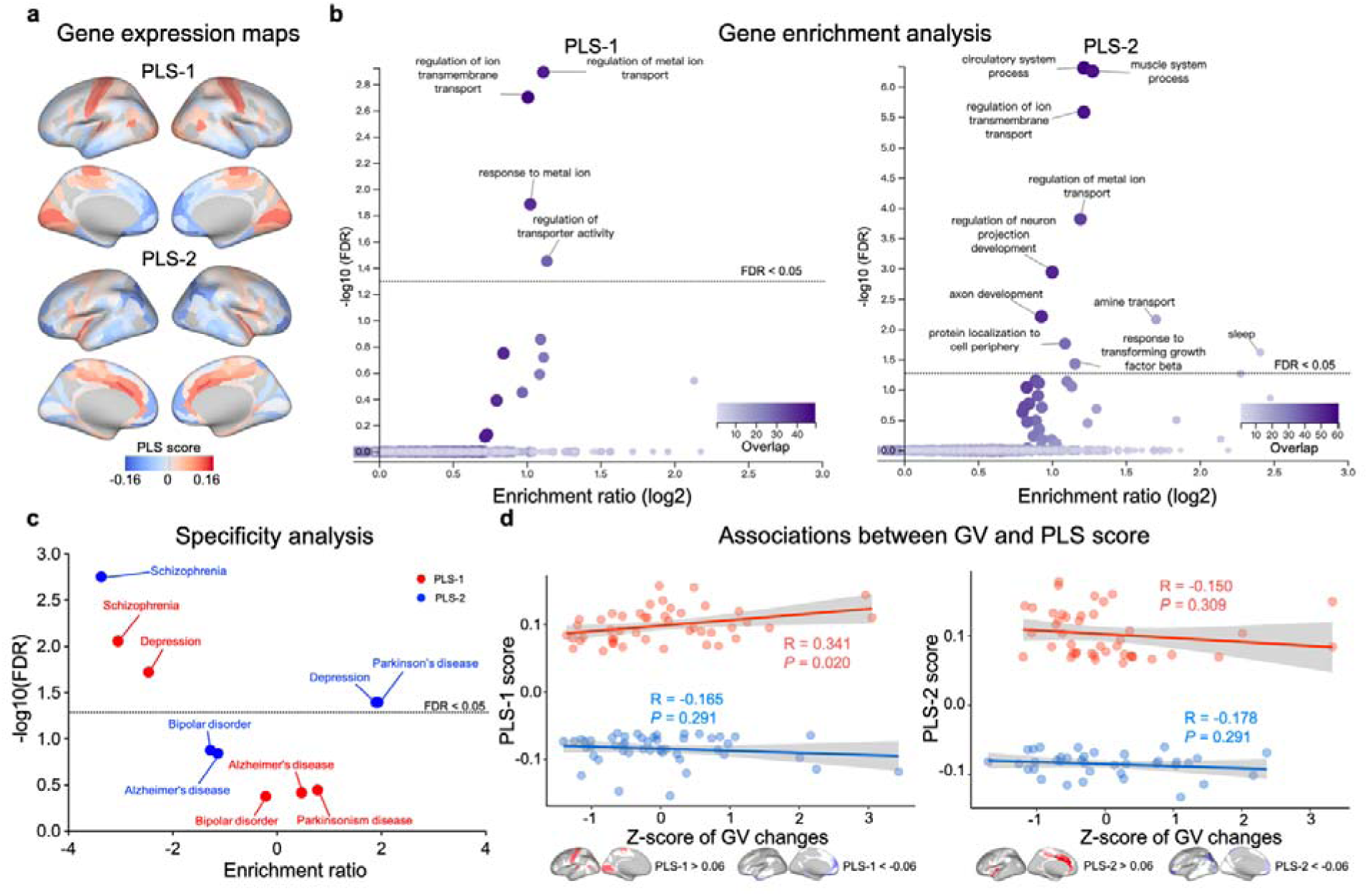
Association between WD-related gradient alterations and gene expression within the cortex. **(a)** Maps of gene expression within the cortex. The color scale indicates the score for PLS-1 and PLS-2, namely the weighted average expression level of 10028. **(b)** Gene enrichment analysis. Genes associated with PLS-1 were enriched for ion transport, ion transmembrane transport, and transporter activity, determining the metal ion homeostasis, and PLS-2 for muscle system process and neuron projection development, as well as ion transport and ion transmembrane transport. In the volcano plots, the x-axis indicates log2 of enrichment ratio and the y-axis indicates-log10 of FDR. Color codes indicate the number of genes related to the biological processes that overlap with the input list of top 10 percentile genes; the dotted line indicates FDR = 0.05. **(c)** Specificity analysis. Concerning risk genes, PLS-1 (red) and PLS-2 (blue) were enriched for genes associated with Schizophrenia, and depression, while PLS-2 was additionally enriched for those associated with Parkinson’s disease. The dotted line indicates FDR = 0.05. **(d)** Associations between GM volume changes and scores higher than 0.06 and lower than −0.06 in both PLS-1 and PLS-2. The PLS-1 scores higher than 0.06 was significantly correlated with GM volume changes. GV, GM volume.

For subcortical regions, two PLS components explained 28% (PLS-1, *P* = 0.0264) and 22% (PLS-2, *P* = 0.0437) of the covariance between the gradient changes of WD and gene expression (**Supplementary Fig. 7**). PSL-1 represented a gene expression profile with high expression mainly in the pallidum and thalamus but low expression in hippocampus and striatum (**Fig. 5a**). PSL-2 revealed a high gene expression in the pallidum, striatum, and amygdala but low expression mainly in the thalamus (**Fig. 5a**). Specificity analysis showed that PLS-1 (**Fig. 5b**)was enriched for the risk gene of WD (PLS-1: *P* = 0.022, ER = −1.946), with PLS-1 and PLS-2 additionally enriched for the gene causing Schizophrenia (PLS-1: FDR = 0.0155, ER = −2.73) and Alzheimer’s disease (PLS-2: FDR = 0.0425, ER = −2.16). The GM volume changes had a significant negative correlation with PLS-1 scores higher than 0.06, indicating that structural impairments and gradient perturbation in subcortical regions also be mediated by the similar transcriptional specializations (**Fig. 5c**).

**Fig. 5.**
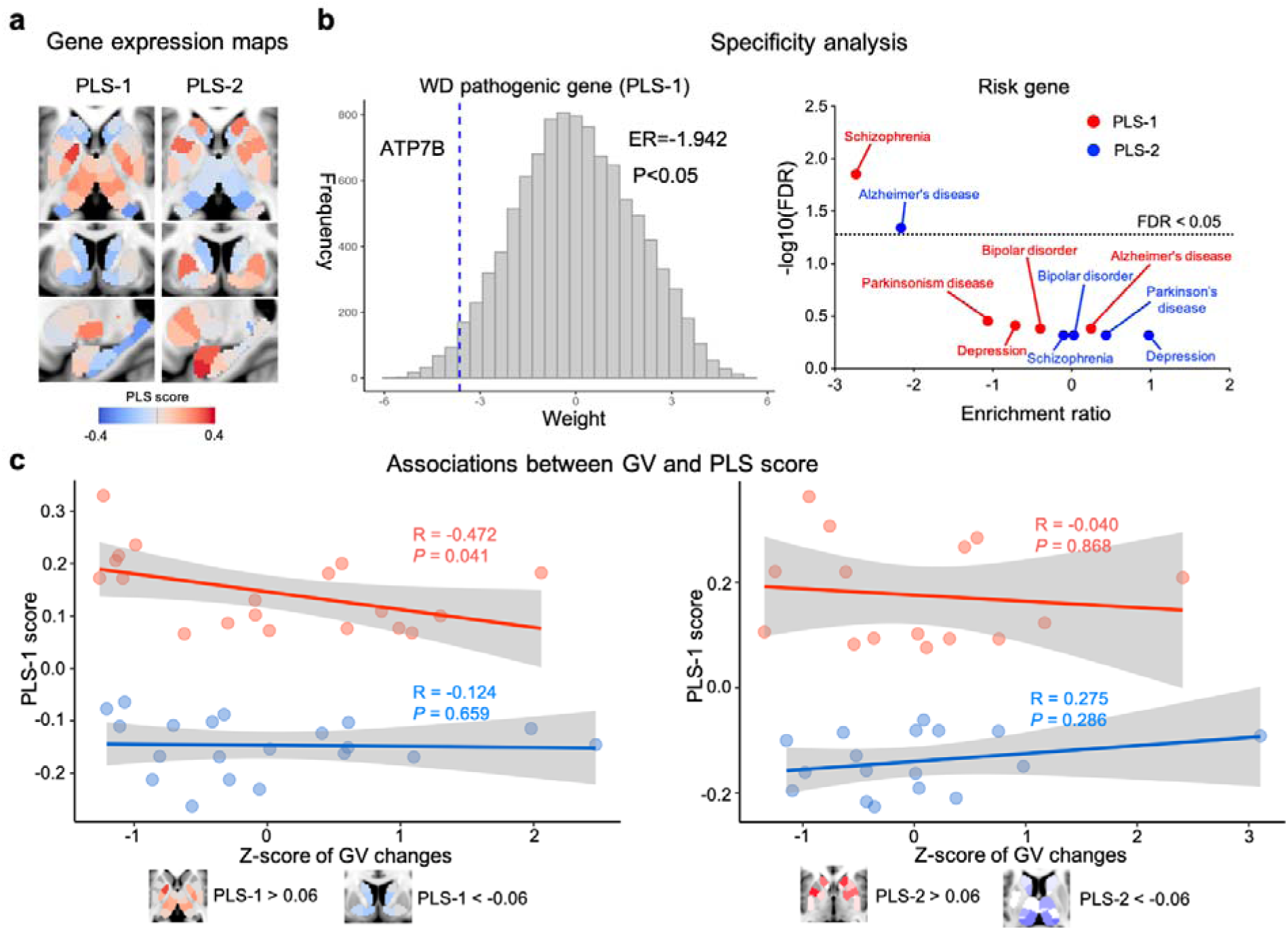
Association between WD-related gradient alterations and gene expression within the subcortical regions. **(a)** Maps of gene expression within the subcortical regions. The color scale indicates the score for PLS-1 and PLS-2, namely the weighted average expression level of 10028. **(b)** Specificity analysis. PLS-1 was significantly enriched for WD pathogenic genes (ATP7B); the histogram shows bootstrap weights of 10000 permutations; the dotted line indicates the bootstrap weight of the candidate genes. Concerning risk genes, PLS-1 (red) was enriched for genes associated with Schizophrenia, while PLS-2 (blue) was enriched for those associated with Alzheimer’s disease. The dotted line indicates FDR = 0.05. GV, GM volume. **(c)** Associations between GM volume changes and scores higher than 0.06 and lower than −0.06 in both PLS-1 and PLS-2. The PLS-1 scores higher than 0.06 was significantly correlated with GM volume changes.

## DISCUSSION

Macroscale hierarchy has been widely regarded as a key principle of brain organization^13^. It has been involved in multiple systems that span from primary to transmodal networks^17^. Hierarchical representations account for the spatial arrangement of local processing streams throughout the cerebral cortex, ensuring sensory signals progressively to integrating as abstract representations^17^. Since WD has been clinically characterized as a disorder with multiple neurological and psychiatric phenotypes^36^, it may result in errors in information flow from sensorimotor to cognitive processing. Accordingly, here we assessed the pattern of functional hierarchy in larger samples of WD and HCs, based on an advanced connectome gradient approach. Both WD and HCs groups revealed that principal functional gradient mapping captured the macroscale axis of connectivity variations with primary network and transmodal DMN anchored at two opposite ends and the remaining networks emerging in-between^17^. Naturally, this gradient of WD was globally suppressed at both ends compared to HCs. Consistent with these results, VBM analysis also illustrated that in WD, GM volume significantly decreased in multiple systems, including SMN, VIS, and transmodal areas. Further spatial correlations between gradient and GM volume revealed function-structure decoupled in VIS, VAN, and SUB, suggesting hierarchical imbalances related to structural alterations. In sum, our multimodal analysis provides converging evidence for that structural vulnerability might shape the aberrant connectome hierarchy.

At the regional level, patients with WD exhibited an extended principal gradient, showing an increase in gradient score in primary systems such as the SMA and V1, and a decrease in transmodal regions including the mTP and mPFC. Generally, an important role of SMA and V1 in motor control and visual processing has been identified in the motor and visual system, particularly for postural stabilization of the body and spatial pattern recognition^37–40^. Consistent with the clinical impairment in these regions, WD-related abnormalities of intrinsic brain activity in the SMN and VN have been highlighted in RS-fMRI^8^ and tomography-electrophysiology studies^41^. Likewise, cortical thickness damage in these regions was also reported to be correlated with the neurological symptoms in WD^42^. DMN areas, such as mPFC and mTP, are thought to be involved in several different functions, such as self-referential, introspective cognition as well as mentalizing^43,44^. Of note, patients with WD have neuropsychological and attention network deficits^9,45^. Moreover, containing these and other nodes in DMN, multiple previous RS-fMRI studies have already reported a series of results indicative of functional connectivity alterations in WD^7^. These results were also confirmed by our meta-analysis, revealing WD-related increased gradient scores in the areas involving low-level sensory and motor-related processes but brain regions with decreased gradient scores mainly involving higher-order cognitive function. Interestingly, we also observed an increased gradient score in unimodal areas, such as the insular, which is located between the primary systems and DMN^17^. Collectively, these findings suggest that the imbalance in the functional hierarchy might arise from the disorder of local processing streams transformed system-by-system in WD cerebral cortex. More notably, our results also showed increasing gradient scores in SUB, such as the putamen and thalamus. The SUB is the most damaged brain structure in WD. Due to the impaired regions leading to disrupted networks^16^, the interaction among SUB and between subcortio-cortical networks could impact the topology of the subcortical macroscale connectome in WD. Again, recent studies have reported that connectivity among SUB was highly associated with deficits in neurological symptoms^46^ while subcortico-cortex was related to neuropsychological disorder in WD^9^. In this context, the relationship between gradient and UWDRS-N was mediated by GM volume alteration, furtherly attesting that structural vulnerability impacts macroscale hierarchy and ultimately leads to deficits in WD phenotype. Taking all the above into consideration, our findings indicate that complex clinical symptoms of WD might arise from perturbation in functional hierarchy across individuals that is not merely due to aberrant spatial patterns, but rather linked to system-level deficits.

In addition to offering a novel perspective on the structural underpinning of perturbation in the functional hierarchy, we incorporated a PLS regression to establish a link between WD-related gradient changes and gene expression. Within the cortex, PLS-1 reflected regulation of gene expression at ion transmembrane transport, metal ion transport, and transporter activity. PLS-2 also mediated the regulation of ion transmembrane transport, but more reflected regulation of neuron projection development and axon development, as well as a muscle system process. In biology, the transporter is a transmembrane protein that moves ions across biological membranes to accomplish different biological functions, such as cellular communication and maintaining homeostasis^47^. Ion transport is a biological process that transports ions against the concentration gradient–from high concentration to low concentration^48^. Neuron projection is any process extending from a neural cell, such as axons and dendrites. its development is the progression from neural formation to the mature structure^49^ and also promotes neuron differentiation^50^. The muscle system, in vertebrates, is controlled through the nervous system, and its impaired leads to different movement disorders^51^. Indeed, WD is a disorder of copper transport within the cell resulting in copper accumulation in the brain^52^, which further triggers neuron degeneration. Therefore, PLS-1 likely represents biological mechanisms underlying ion homeostasis inside and outside nerve cells. On the other hand, PLS-2 was more associated with neural organization and cell growth as well as controlling muscle signaling. The specificity analysis exhibited that PLS-1 and PLS-2 were both associated with risk genes for schizophrenia and depression and that PLS-2 was additionally associated with genes causing Parkinson’s disease. Although no association between WD pathogenic gene (ATP7B) and PLS components was observed within the cortex, our results explained why WD patients present multiple complex neurological and psychiatric symptoms at the biological level.

More importantly, within subcortical regions, we demonstrated that PLS-1 was associated with WD pathogenic gene, which indicates that the ATP7B gene has lower expression in subcortical regions. The present work is the first to provide evidence for the role of the WD pathogenic gene impacting subcortical function, which is also attested by an animal model study, revealing that dysfunctional ATP7B protein is related to the local copper accumulation of SUB and the manifestations in WD^4^. The relevance of these PLS components with other risk genes was also calculated by specificity analysis, showing that PLS-1 was associated with risk genes of schizophrenia while PLS-2 was related to genes causing Alzheimer’s disease. Consistent with our results, recent studies have reported that the risk genes of schizophrenia^53^ and Alzheimer’s disease^54^ are significantly associated with subcortical dysfunction. Interestingly, we found that the spatial pattern of GM volume alteration is similar to that of PLS-1 with higher expression in subcortical regions, and this association was also exhibited in the cerebral cortex. These findings indicate that the GM volume impairments and gradient perturbation are mediated by similar transcriptional specializations.

Findings have to be interpreted in the context of potential limitations. Gene set from AHBA were sampled from donors without a diagnosis of WD. Moreover, the associations between connectome gradient and gene expression were susceptible to intersubject variability. Likewise, the group-averaged gradient was associated with gene expression data, which could not provide a powerful genetic biomarker for individual prediction. In the future, a larger sample of whole-brain gene expression data from WD patients is required to further represent and validate the relationship between connectome gradient and transcriptome. Future studies should also add within-subject design to perform individual-level prediction for WD.

This study revealed the gradient perturbations related to cognitive terms and clinical phenotypes in WD. Then, we mapped associations of these gradient differences with specific GM volume alterations and transcriptional expressions. Further, we uncovered the biological underpinnings of these gradient-derived gene sets by enrichment analysis. The biological underpinnings was determined by underlying factors including 1) function-structure decoupled characterized by spatial correlation alterations in WD; 2) high associations between gene expression and gradient-differences and identified biological process within the cerebral cortex; 3) implications in psychiatric and neurological diseases and for the first time characterizing the role of ATP7B impacting subcortical function; 4) GM volume impairments and perturbation of connectome gradients are mediated by the similar transcriptional specializations. In sum, these findings mapping the structural and transcriptomic underpinnings of gradient perturbation in WD provide a deep insight into WD neurobiological underpinnings underlying the emergence of complex neurological and psychiatric phenotypes.

## Data Availability

Anonymised data are available on request to the corresponding author. Gene expression data that was used for transcriptional analysis can be found in the ABHA database (https://human.brainmap.org/static/download).

## ACKNOWLEDGEMENT

This work was supported by the National Natural Science Foundation of China (81973825 and U22A20366), the University Synergy Innovation Program of Anhui Province (GXXT-2020-025), the Project BEBPC-TCM (2019XZZX-NB001), the Natural Science Research Project of Anhui Universities (KJ2021A0580 and KJ2021A0555), the Natural Science Foundation of Anhui Province (2108085QH367) and the Open Fund Project of Key Laboratory of Xin’an Medicine of Ministry of Education (2020xayx12).

## AUTHOR CONTRIBUTION

W.Y and BQ contributed to the study concept and design; S.H contributed to Conceptualization, Methodology, Software, Visualization, and Writing -Original Draft; C.L, Y.W and T.W contributed to Writing - Review & Editing and Methodology; X.W contributed to Review & Editing; T.D, Y.Y and Y.D contributed to the acquisition the data.

