## Supplementary Materials for "Functional gradient perturbation in Wilson disease correlates with structural lesions and transcriptomic specializations"

*** Corresponding at:**

Wenming Yang or Bensheng Qiu, Department of Neurology, The First Affiliated Hospital of Anhui University of Chinese Medicine, 230031 P.R. China; or Center for Biomedical Imaging, University of Science and Technology of China; Hefei, Anhui, 2300026 P.R. China.

**SI Materials and Methods**

**fMRI data preprocessing**

Preprocessing of the functional images involved the following steps: (1) discarding the first 10 time points of functional images to account for magnetic stabilization; (2) slice timing correction to compensate for temporal shifts of different slices; (3) within-subject fMRI image realignment to estimate and spatially correct for head motions of different volumes; (4) rigid-body T1 image registration to the functional mean image and functional image normalization to the Montreal Neurological Institute (MNI) standard space using the T1 images; (5) functional image resampling to 3×3×3 mm^3^ voxel size followed by regressing out confounding signals, including linear trends, white matter (WM), cerebrospinal fluid (CSF), and Friston’s 24 head motion parameters; (6) bandpass filtering (0.01-0.1 Hz) to suppress low-frequency drift and physiological noises (breathing and heartbeat); and (7) functional image spatial smoothing using a 6 mm full-width at half-maximum (FWHM) Gaussian kernel. All data preprocessing was performed on DPABI version 5.3 (http://rfmri.org/dpabi), a MATLAB toolbox for batch fMRI data preprocessing.

**Supplementary Figures**

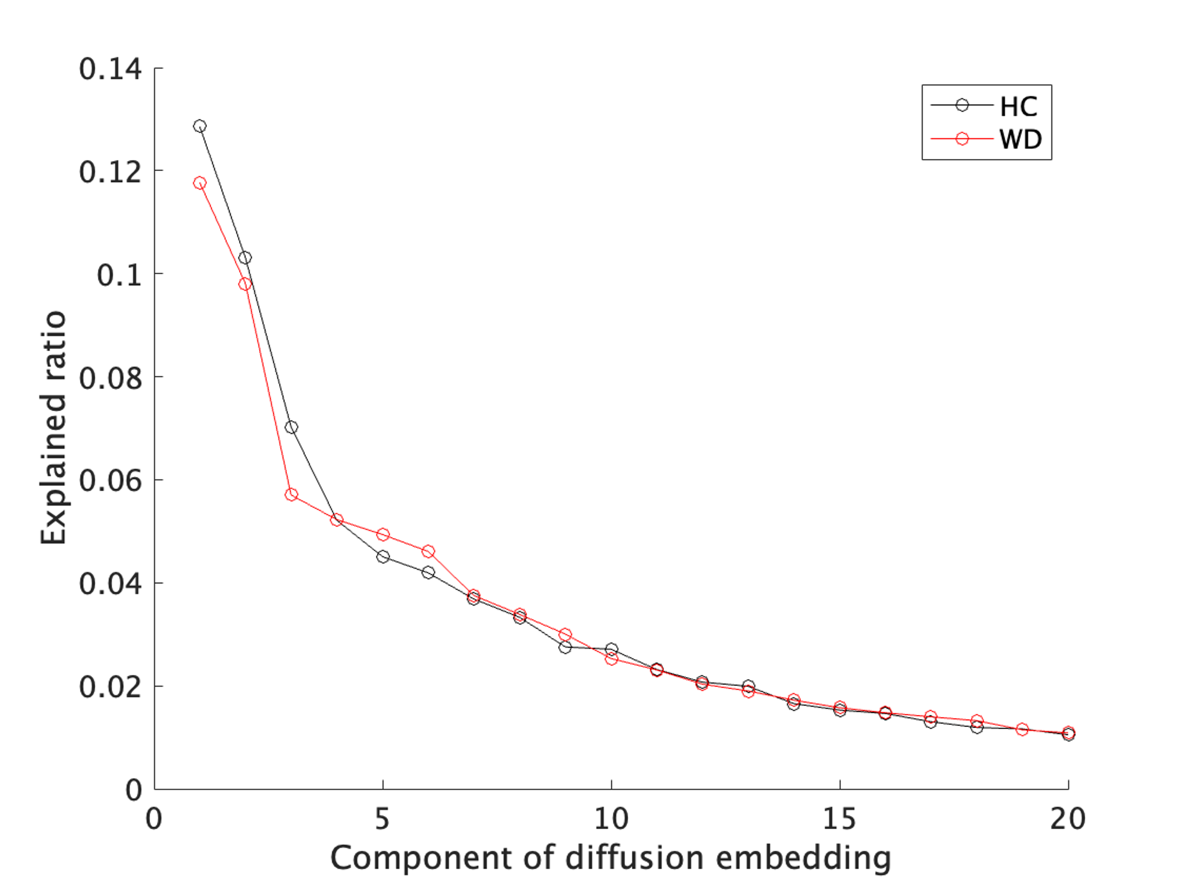

**Supplementary Fig. 1** The averaged explained ratio (λ values). The first two gradients explained 22.2 ± 2.9% of the total variance in the connectome across all individuals (WD: 21.6 ± 2.9%, HC: 23.2 ± 2.8%).

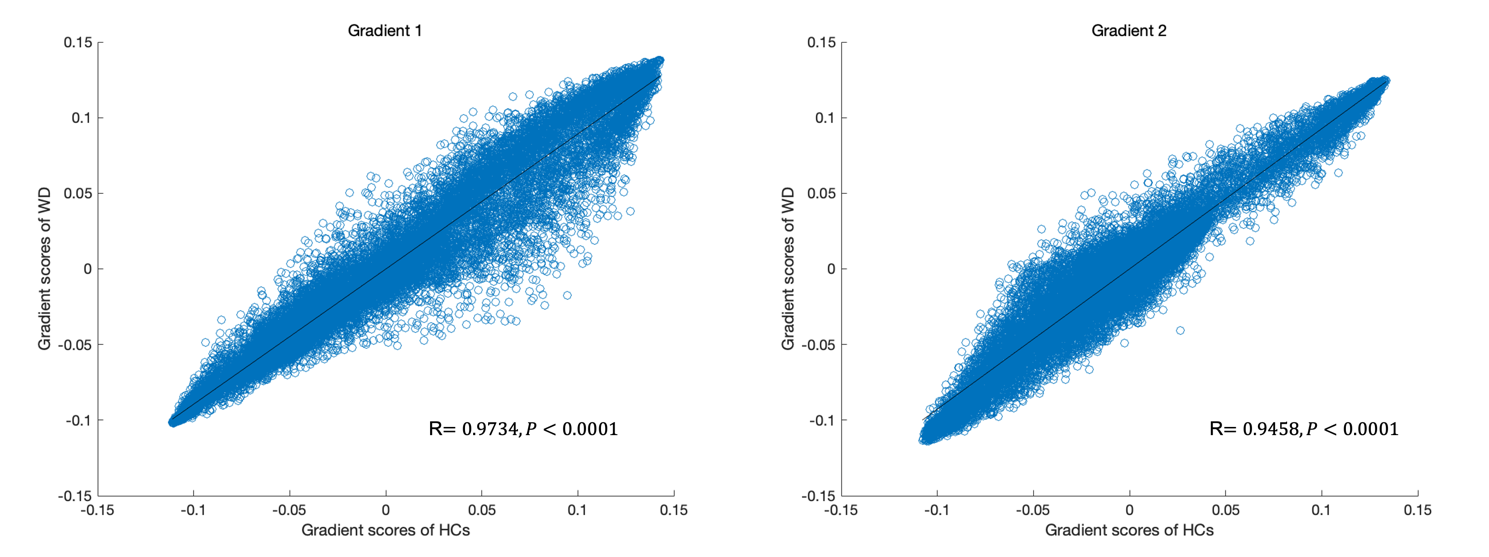

**Supplementary Fig. 2** Spatial correlations of the group-averaged gradient maps between the WD and HCs. These correlations were corrected for spatial autocorrelations by using a permutation test (N = 10,000).

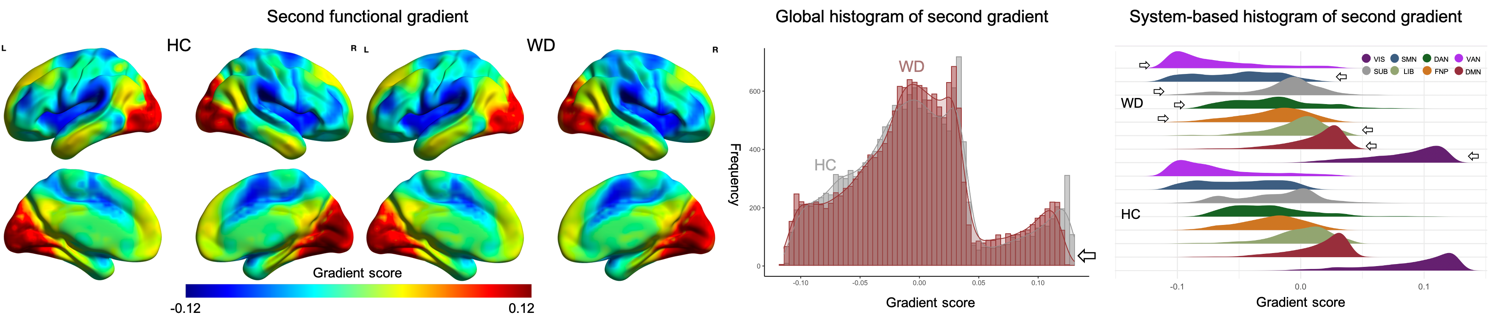

**Supplementary Fig. 3** Secondary gradient mapping in patients with MDD and controls. The global and network-based histograms show that the extreme values in the patients with WD were contracted relative to those in the HCs in the secondary gradient. VIS, visual network; SMN, sensorimotor network; DAN, dorsal attention network; VAN, ventral attention network; SUB, subcortical regions; LIB, limbic network; FPN, frontoparietal network; DMN, default mode network.

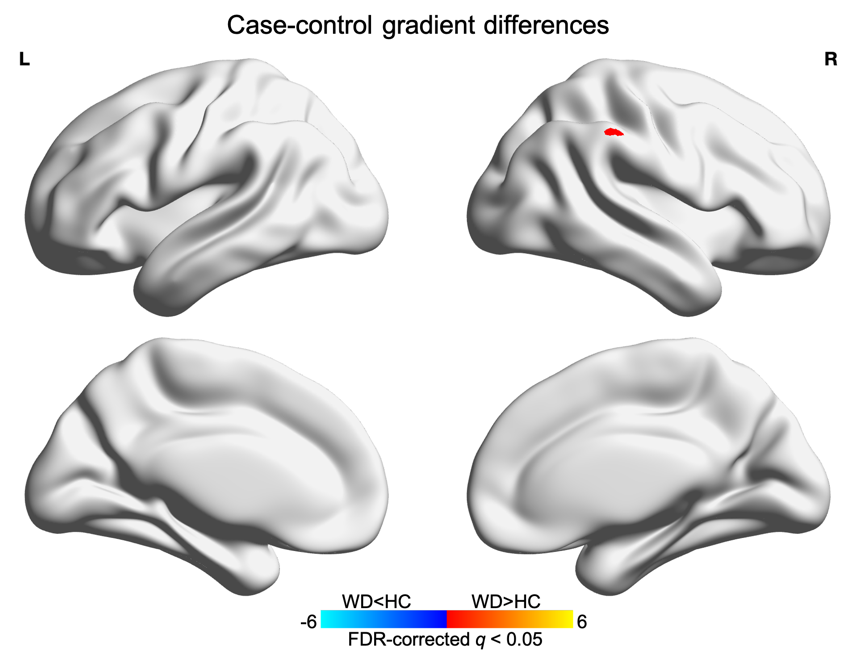

**Supplementary Fig. 4** Statistical comparison of the secondary gradients between the WD and HCs.

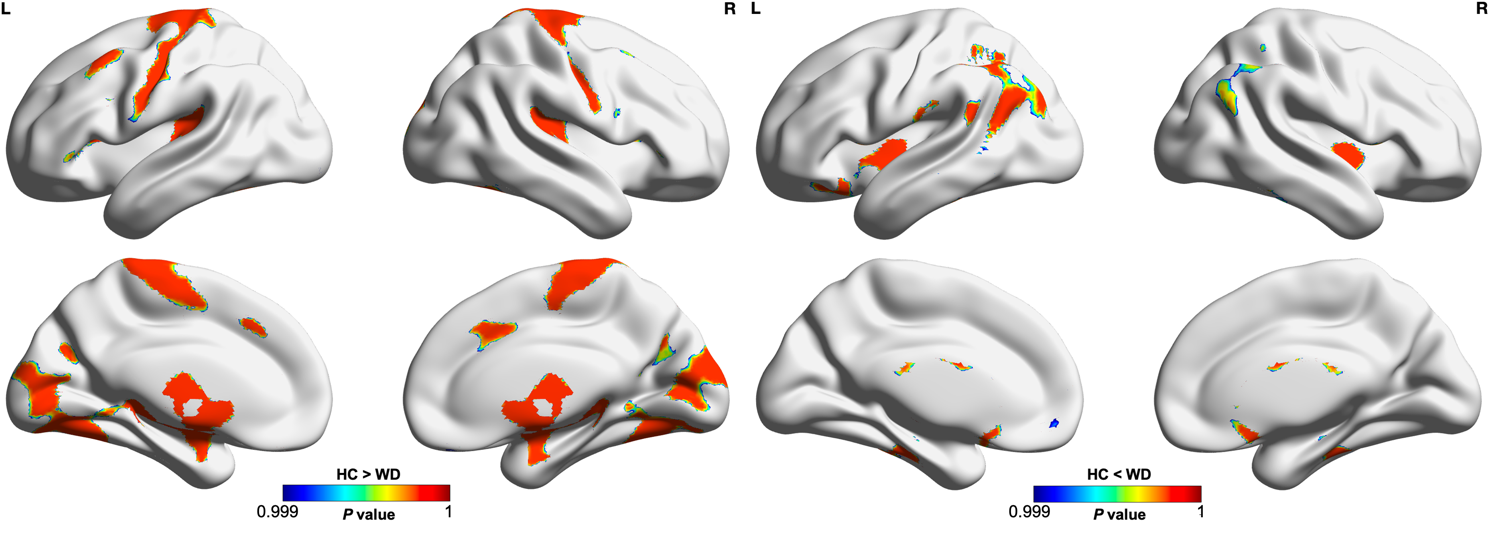

**Supplementary Fig. 5** The group differences of GM volume between WD and HCs.

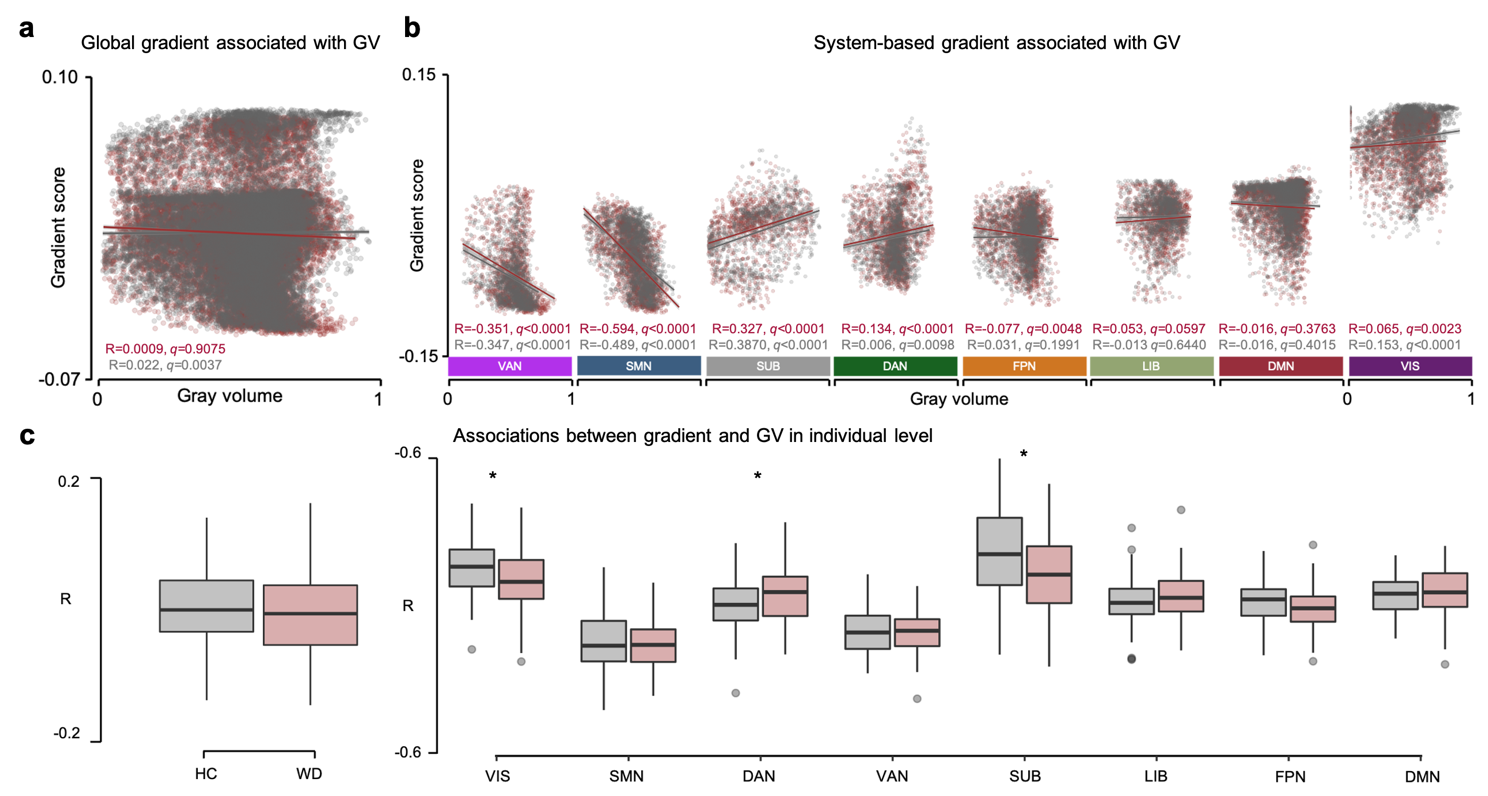

**Supplementary Fig. 6** Spatial correlations between secondary gradient and GM volume. **a.** Spatial correlation between global secondary gradient and GM volume. **b.** Network-based spatial correlations between secondary gradient and GM volume. **c.** The differences of global and network-based spatial correlations between WD and HCs across subjects. *, *P* < 0.05. VIS, visual network; SMN, sensorimotor network; DAN, dorsal attention network; VAN, ventral attention network; SUB, subcortical regions; LIB, limbic network; FPN, frontoparietal network; DMN, default mode network.

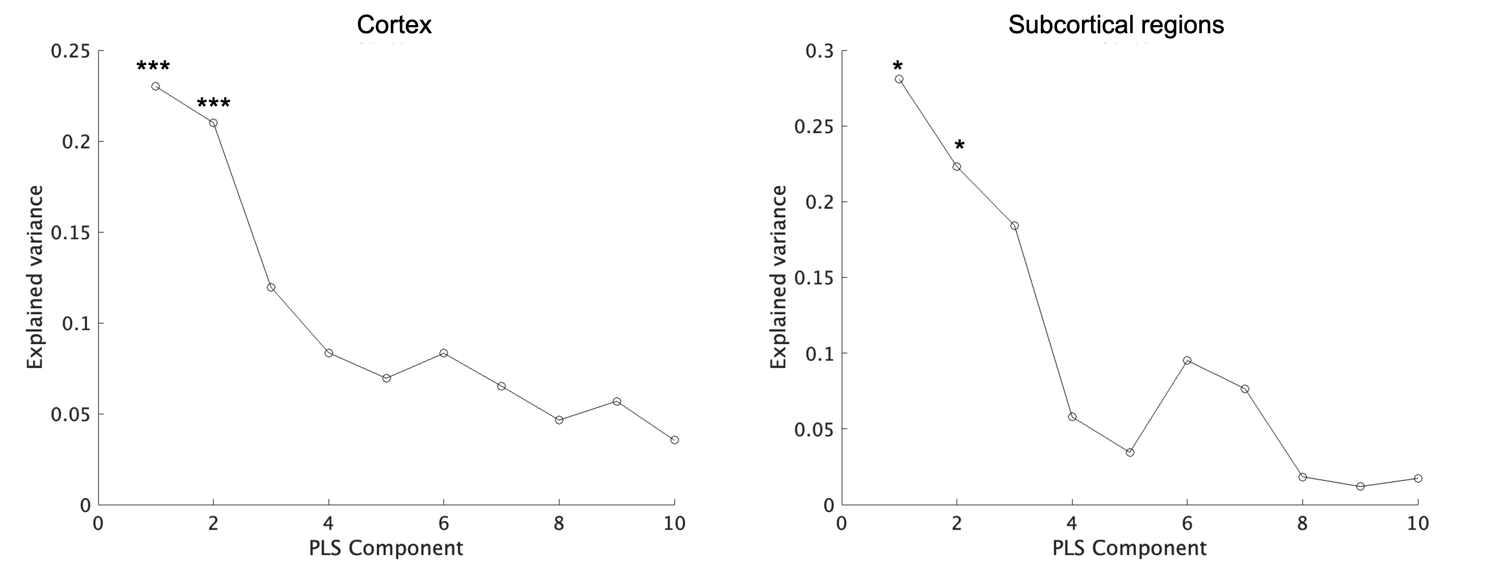

**Supplementary Fig. 7** The percentage of variance in the response variables explained by the components in the partial least squares regression analysis. The significance level was determined by a permutation test (N = 10,000) with spatial autocorrelation corrected. ***, *P* <0 .001; *, *P* < 0.05.

**Supplementary Tables**

**Supplementary Table 1.** The lists of candidate genes used for disease specificity analysis are presented in Fig. 4 and 5.

| A | A2M, ACE, ACHE, APBA1, APBB2, APLP1, APLP2, APOC1, APP, BACE2, BCHE, BLMH, CASP3, CHRNA3, CTSB, DBN1, ESR1, GSK3B, IL1B, KCNIP3, KLK6, LRP1, LRRC15, MAPT, PLAU, PSEN1, PSEN2, SORL1 |
| --- | --- |
| B | ATP1A3, DBH, DDC, HTRA2, LRRK2, NR4A2, PARK2, PARK7, PINK1, SLC6A3, SNCA, SNCAIP, SNCB, TH, UCHL1 |
| C | ACTN2, AHI1, AKT1, ANKHD1, APOL1, APOL2, APOL4, ATF4, ATF5, ATF7IP, BDNF, CCDC141, CHRM1, CHRNA7, CIT, COMT, DAO, DAOA, DISC1, DRD1, DRD3, DRD5, DTNBP1, ERBB3, ERBB4, FEZ1, GABRA6, GABRB2, GAD1, GRIN1, GRIN2A, GRIN2B, GRM2, GRM3, GRM7, HOMER1, HTR2A, HTR2C, HTR6, ITSN1, KCNN3, MAP1A, NDE1, NDEL1, NPAS3, NRG1, PAFAH1B1, PCNT, PDE4B, PDLIM5, PLXNA2, PPP1R1B, PPP3CA, PPP3CC, PRODH, PVALB, RANBP9, RASSF7, RGS4, RTN4, RTN4R, SLC17A7, SPTBN4, SYN2, SYNE1, TAAR6, TNF |
| D | ADRA2A, AVPR1B, CHRM2, CNR1, CREB1, CRH, CRHR1, CRHR2, CUX2, GAD2, GPR50, HTR1A, HTR1B, HTR1D, HTR3A, HTR5A, MAOA, PDE1A, SLC6A2, SLC6A4, SST, TAC1, TPH1, TPH2 |
| E | ADRBK2, CLOCK, XBP1 |

(A) Alzheimer’s disease, (B) Parkinson‘s disease, (C) Schizophrenia, (D) Depression, (E) Bipolar disease

**Supplementary Table 2.** Between-group differences in the principal gradient scores of the subnetworks in the mean maps

| Subnetworks | *t* | Cohen’s *d* | *P* | *FDR q* |
| --- | --- | --- | --- | --- |
| Visual network | 38.475 | 0.777 | <1.0×10^-20^ | <1.0×10^-20^ |
| Sensorimotor network | 45.071 | 0.970 | <1.0×10^-20^ | <1.0×10^-20^ |
| Dorsal attention network | 1.889 | 0.046 | 0.0589 | 0.0589 |
| Ventral attention network | 20.223 | 0.523 | <1.0×10^-20^ | <1.0×10^-20^ |
| Limbic network | -25.348 | 0.699 | <1.0×10^-20^ | <1.0×10^-20^ |
| Frontoparietal network | 9.3046 | -0.201 | 3.221×10^-20^ | 4.294×10^-20^ |
| Default mode network | -29.036 | -0.503 | <1.0×10^-20^ | <1.0×10^-20^ |
| Subcortical network | 8.032 | 0.254 | 2.685×10^-15^ | 3.069×10^-15^ |

Abbreviations: FDR, false discovery rate.

**Supplementary Table 3** Clusters with significant between-group differences in the principal primary-to-transmodal gradient

| No. | Region | x | y | z | Z (Peak) | Cohen’s *d* | Size (voxel) |
| --- | --- | --- | --- | --- | --- | --- | --- |
| WD > HCs | |  |  |  |  |  |  |
| 1 | Right SMA | 2 | -10 | 60 | 4.91 | 0.13 | 128 |
| 2 | Right primary visual cortex | 10 | -86 | 8 | 4.27 | 0.10 | 126 |
| 3 | Left putamen | -18 | 14 | -4 | 5.58 | 0.17 | 61 |
| 4 | Left insular lobe | -34 | 22 | 8 | 4.64 | 0.12 | 57 |
| 5 | Right putamen | 22 | 10 | -4 | 5.81 | 0.18 | 49 |
| 6 | Left middle frontal gyrus | -26 | 38 | 24 | 4.94 | 0.13 | 42 |
| 7 | Right inferior temporal gyrus | 50 | -62 | -8 | 3.87 | 0.08 | 35 |
| WD < HCs | |  |  |  |  |  |  |
| 8 | Right medial temporal pole | 38 | 14 | -40 | -5.75 | -0.18 | 217 |
| 9 | Left temporal pole | -34 | 18 | -24 | -5.47 | -0.16 | 210 |
| 10 | Left superior frontal gyrus | -22 | 46 | 44 | -5.22 | -0.15 | 160 |
| 11 | Right medial prefrontal cortex | 10 | 50 | 48 | -5.83 | -0.18 | 139 |
| 12 | Left rectal gyrus | -6 | 54 | -24 | -5.92 | -0.19 | 52 |

**Supplementary Table 4.** Between-group differences in the secondary gradient scores of the subnetworks in the mean maps

| Subnetworks | *t* | Cohen’s *d* | *P* | *FDR q* |
| --- | --- | --- | --- | --- |
| Visual network | -29.634 | -0.598 | <1.0×10^-20^ | <1.0×10^-20^ |
| Sensorimotor network | -7.230 | -0.156 | 6.674×10^-13^ | 7.628×10^-13^ |
| Dorsal attention network | 10.824 | 0.265 | <1.0×10^-20^ | <1.0×10^-20^ |
| Ventral attention network | 11.796 | 0.305 | <1.0×10^-20^ | <1.0×10^-20^ |
| Limbic network | -9.702 | -0.267 | <1.0×10^-20^ | <1.0×10^-20^ |
| Frontoparietal network | 12.446 | 0.269 | 3.221×10^-20^ | 4.294×10^-20^ |
| Default mode network | -19.399 | -0.336 | <1.0×10^-20^ | <1.0×10^-20^ |
| Subcortical network | 4.758 | 0.151 | 2.241×10^-6^ | 2.241×10^-6^ |

Abbreviations: FDR, false discovery rate.

**Supplementary Table 5** Clusters with significant between-group differences in the secondary gradient

| No. | Region | x | y | z | *T* | Cohen’s *d* | Size (voxel) |
| --- | --- | --- | --- | --- | --- | --- | --- |
| WD > HCs | |  |  |  |  |  |  |
| 1 | Right supramarginal gyrus | 66 | -30 | 44 | 6.00 | 0.13 | 0.19 |

**Supplementary Table 6.** Spatial correlation between the meta-analytic map of cognitive terms and WD-related alterations in the primary gradient

| WD-positive | |  | WD-negative | | |  |
| --- | --- | --- | --- | --- | --- | --- |
| Term | | *r* | | Term | | *r* |
| motor | 0.253 | | | | theory mind | 0.258 |
| somatosensory | 0.228 | | | | mind | 0.24 |
| movement | 0.216 | | | | social | 0.235 |
| movements | 0.201 | | | | mental states | 0.231 |
| pain | 0.196 | | | | mind tom | 0.222 |
| motor imagery | 0.189 | | | | mentalizing | 0.207 |
| hand | 0.196 | | | | theory | 0.187 |
| painful | 0.188 | | | | person | 0.182 |
| execution | 0.186 | | | | autobiographical memory | 0.152 |
| finger | 0.165 | | | | beliefs | 0.135 |
| visual | 0.154 | | | | construction | 0.123 |
| gain | 0.14 | | | | episodic | 0.121 |
| imagery | 0.139 | | | | social cognition | 0.121 |
| foot | 0.138 | | | | memories | 0.12 |
| primary visual | 0.136 | | | | dementia | 0.118 |
| early visual | 0.135 | | | | self-referential | 0.117 |
| action | 0.131 | | | | thinking | 0.094 |
| preparation | 0.129 | | | | memory retrieval | 0.094 |
| sighted | 0.123 | | | | comprehension | 0.093 |
| extrastriate | 0.122 | | | | episodic memory | 0.091 |
| finger movements | 0.116 | | | | semantic memory | 0.091 |
| vision | 0.116 | | | | social interaction | 0.085 |
| motor task | 0.116 | | | | intensions | 0.082 |
| sensory | 0.115 | | | | experiences | 0.081 |
| actions | 0.111 | | | | rest | 0.069 |
| tapping | 0.11 | | | | sentence | 0.068 |
| executed | 0.108 | | | | read | 0.059 |
| mirror | 0.107 | | | | concrete | 0.057 |
| force | 0.095 | | | | reasoning | 0.057 |
| stimulation | 0.094 | | | | judgments | 0.054 |

**Supplementary Table 7** Clusters with significant between-group differences in GM volume

| Subnetworks | WD < HC | | WD > HC | |
| --- | --- | --- | --- | --- |
|  | *P* (Mean) | Voxel size | *P* (Mean) | *Voxel size* |
| Visual network | 0.9997 | 5283 | 0.9997 | 747 |
| Sensorimotor network | 0.9997 | 6793 | 0.9997 | 450 |
| Dorsal attention network | 0.9996 | 1273 | 0.9997 | 1676 |
| Ventral attention network | 0.9996 | 1250 | 0.9997 | 1928 |
| Limbic network | 0.9993 | 184 | 0.9997 | 1523 |
| Frontoparietal network | 0.9996 | 1010 | 0.9997 | 1572 |
| Default mode network | 0.9996 | 5283 | 0.9997 | 747 |
| Subcortical network | 0.9998 | 4434 | 0.9997 | 269 |
